# Long-term predictions of humoral immunity after two doses of BNT162b2 and mRNA-1273 vaccines based on dosage, age and sex

**DOI:** 10.1101/2021.10.13.21264957

**Authors:** Chapin S. Korosec, Suzan Farhang-Sardroodi, David W. Dick, Samaneh Gholami, Mohammad Sajjad Ghaemi, Iain R. Moyles, Morgan Craig, Hsu Kiang Ooi, Jane M. Heffernan

## Abstract

**Summary:** *Background:* The lipid nanoparticle (LNP)-formulated mRNA vaccines are a widely adopted two-dose vaccination public health strategy to manage the COVID-19 pandemic. Clinical trial data has described the immunogeneicity of the vaccine, albeit within a limited study time frame. Our aims were to use a within-host mathematical model for LNP-formulated mRNA vaccines, informed by available clinical trial data, to project a longer term understanding of humoral immunity as a function of vaccine type, dosage amount, age, and sex.

*Methods:* We developed a mathematical model describing the immunization process of LNP-formulated mRNA vaccines, and fit our model to twenty-two clinical humoral and cytokine BNT162b2 or mRNA-1273 human two-dose vaccination data sets. We incorporated multi-dose effects in our model to specify whether the dosage is standard or low-dose. We further specify the age groups 18-55, 56-70, and 70+ in our fits for two-standard doses of mRNA-1273, and sex in our fits for two-standard doses of BNT162b2. We used non-linear mixed effect models to fit to all similar data types (e.g. standard two-dose BNT162b2 or mRNA-1273, or two low-dose mRNA-1273). Therefore, in our fits all estimated parameters are statistically correlated, which allowed us to determine the underlying ‘population-dynamics’ structure common to a data type. We therefore made accurate long-term predictions informed by all clinical data used in this study.

*Findings:* We estimate that two standard doses of either mRNA-1273 or BNT162b2, with dosage times separated by the company-mandated intervals, results in individuals loosing more than 99% humoral immunity relative to peak immunity by eight months following the second dose. We predict that within an eight month period following dose two (corresponding to the CDC time-frame for administration of a third dose), there exists a period of time longer than one month where an individual has less then 99% humoral immunity relative to peak immunity, regardless of which vaccine was administered. We further find that age has a strong influence in maintaining humoral immunity; by eight months following dose two we predict that individuals aged 18-55 have a four-fold humoral advantage compared to aged 56-70 and 70+ individuals. We find that sex has little effect on the vaccine uptake and long-term IgG counts. Finally, we find that humoral immunity generated from two low doses of mRNA-1273 decays substantially slower relative to peak immunity gained than compared to two standard doses of either mRNA-1273 or BNT162b2.

*Interpretation:* For the two dose mRNA vaccines, our predictions highlight the importance of the recommended third booster dose in order to maintain elevated levels of antibodies. We further show that age plays a critical role in determining the antibody levels. Hence, a third booster dose may confer an immuno-protective advantage in older individuals.

*Funding:* This research is supported by NSERC Discovery Grant (RGPIN-2018-04546), NSERC COVID-19 Alliance Grant ALLRP 554923-20, CIHR-Fields COVID Immunity Task Force, NRC Pandemic Response Challenge Program Grant No. PR016-1.

## 1. Introduction

The severe acute respiratory syndrome coronavirus 2 (SARS-CoV-2) first detected in December of 2019 has driven COVID-19 into a global pandemic [1, 2, 3]. Recognizing the severity of this novel pneumonia outbreak, global research efforts were rapidly organized and the first viral sequence became publicly available on 26 December 2019 (LR757995, LR757998) [4]. Immediately, various vaccine research groups began optimizing their current vaccine technologies to express the SARS-CoV-2 protein. SARS-CoV-2 vaccine candidates then resulted, including inactivated virus, viral protein subunits, messenger RNA (mRNA) recombinant human adenovirus, and non-viral replicating vector vaccines. Among these vaccine candidates, mRNA vaccines had a head start in exploiting the viral sequence due to its inherent rapid prototyping (high yield in-vitro transcription reactions) and manufacturing scalability [5]. The pre-clinical trial was initiated immediately in January 2020 and in April 2020, Phase I/II of the clinical trials commenced. In less than 8 months, the mRNA vaccines BNT162b2 (manufactured by Pfizer-BioNTech) and mRNA-1273 (manufactured by Moderna) were approved for emergency use in several countries [6, 7].

To date, 78.54% of the 12 and older population in Canada are fully vaccinated with Pfizer-BioNTech and Moderna comprising 32.38% (12.3 million people) and 9.25% (3.5 million people) of vaccinated individuals, respectively [8]. Further, more than 222 and 150 million doses of Pfizer-BioNTech and Moderna vaccines, respectively, have been administered in the United States [9]. On July 29th, 2021, the Ministry of Health of Israel announced a third-booster-dose strategy based on preliminary report finding vaccine efficacy in prevention of infection via the Pfizer-BioNTech vaccine drops from 75% to 16% seven months following the second dose [10]. There is therefore great urgency to understand and accurately predict waning immunity amongst individuals who received two-doses of the LNP-formulated mRNA vaccines, and to provide an estimate for a correlate of protection.

For acute SARS-CoV-2 infection, antibody and memory B-cell responses have been reported to be robust for up to 8 months [11]. In comparison, two doses of mRNA-1273 and BNT162b2 have shown seroconversion in the short term that surpasses that of recovered patients: one month post second dose, antibody stimulation (IgG and IgM) is reported to be higher while neutralizing antibodies are shown to be similar to recovered patients for fully vaccinated individuals [12, 13, 14]. In vaccinated individuals, studies of antibodies responses are predominantly short- (less than 60 days) and medium-term (less than 120 days) studies [11, 15, 16, 17, 18, 19, 20, 21, 22], with one recent study examining antibody response out to day 210 [23]. With the continued emergence of new variants of concern, there is now a key gap in the understanding of the long-term (beyond 120 days) robustness of the two-dose LNP-formulated mRNA vaccines adopted by health authorities. To address this gap, we established an ODE-based mechanistic model that describes the humoral immune response of mRNA vaccines to predict long term immunity. We fit our model to reported antibody and cytokine levels to clinical trial data using two standard doses of mRNA-1273 (100*μ*g) or BNT162b2 (30*μ*g) [17, 18, 19, 20, 21, 22], as well as separately to two low doses of mRNA-1273 (25*μ*g) [24]. Model results show significant decay in antibody a few months after dose-two inoculation, with the decay rate depending on the vaccine type, dose size, and age of the vaccine recipient. We find little difference between male and female predicted vaccine response.

## 2. Methods

### 2.1. Clinical data acquisition

All clinical data used in this work were previously published and are summarized in Table 1. If unavailable directly from the published source, we digitized the data directly using the software WebPlotDigitizer (version 4.5) [25].

**Table 1:** Summary of clinical data used in this work. Some publications listed studied in-host vaccine dynamics from both Sars-CoV-2 naive and previously infected indivduals, in this work we only digitized those data that are in the naive category and report the associated population size.

| Reference | Quantities used<br>in this work | Vaccine | Dosage | Population<br>size |
| --- | --- | --- | --- | --- |
| [17] | RBD IgG, Spike IgG | BNT162b2 &<br>mRNA-1273 | 2 dose, 30μg<br>or 100μg | 33 (32 BNT162b2 & 1<br>mRNA-1273) |
| [18] | Spike IgG | BNT162b2 | 2 dose, 30 μg | 124 for dose 1<br>69 for dose 2 |
| [19] | Spike-RBD IgG, IFN-γ,<br>IL-6, IL-8, IL-15 | BNT162b2 | 2 dose, 30 μg | 63 |
| [20] | Spike IgG, IFN-γ | BNT162b2 | 2 dose, 30 μg | 20 |
| [21] | RBD IgG | mRNA-1273 | 2 dose, 100 μg | 34 |
| [26] | Spike IgG, RBD IgG | mRNA-1273 | 2 dose, 25 μg | 33 |
| [22] | Spike IgG, RBD IgG | BNT162b2 &<br>mRNA-1273 | 2 dose, 30μg<br>or 100μg | 20 (6 BNT162b2 & 14<br>mRNA-1273) |
| [23] | Spike IgG | BNT162b2 | 2 dose, 30 μg | 46 (24 females, 22 males) |

### 2.2. In-host model for LNP-formulated mRNA vaccination

For this work, we developed a novel in-host mathematical model to describe the vaccination process from lipid nanoparticle-formulated mRNA vaccines, such as those currently in public use by Pfizer (BNT162b2) and Moderna (mRNA-1273). We modelled the time dependence of eight state variables: lipid nanoparticles (*L*), vaccinated cells (*V*), CD4^+^ T cells (*T*), plasma B cells (*B*), antibody (*A*), CD8^+^ T cells (*C*), and the cytokines interferon (*F*) and interleukin (*I*). The model developed in this work is adapted from our recently published adenovirus-based model in order to describe LNP-formulated mRNA vaccines [27]. A detailed description and motivation of our model can be found in the supplemental text, where a summarized description of the state variables, units, and initial conditions can be found in Table S1, and a summarized description of all parameters can be found in Table S2. Predictive simulations based on the Monolix-determined fit parameters were completed in **R** (Version 3.6.2) using the *deSolve* library (Version 1.29), where the parameter values were determined as described in section 2.3.

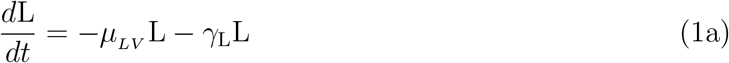

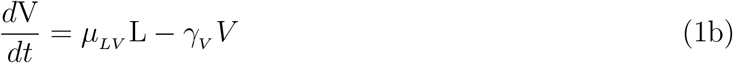

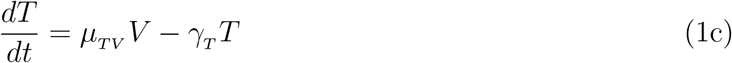

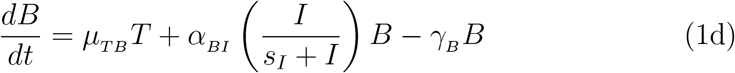

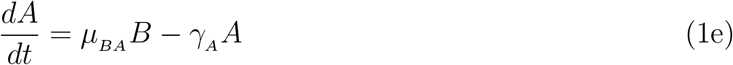

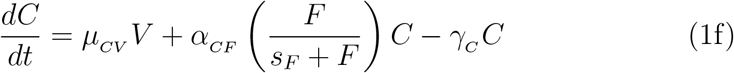

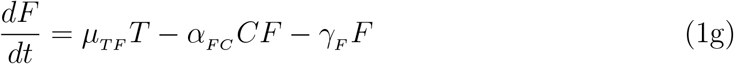

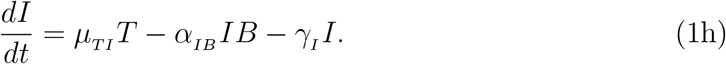

### 2.3. Parameter estimation, fitting assessment, and long-term simulations

All fits to clinical data using our model (Eq. 1) were performed in Monolix [28] (Version 2020R1) using non-linear mixed-effects models. IgG and IFN *− γ* concentrations vary over many orders of magnitude over time, as such we log-transformed these quantities during fitting, while we fit to the linear interleukin response. Individual parameters (Tables S3 and S4) for each data set are determined by the maximum likelihood estimator Stochastic Approximation Expectation–Maximization (SAEM), and all fits met the standard convergence criteria (complete likelihood estimator). For IgG data sets from Ref. [22], which begin on day ∼ 50 past dose one, an interval-censored data point was added to day 1 to guide the initial condition fit, where the allowable initial range of the fit is bounded within all the initial day 1 IgG values used in this work. Final clinical data points for IL-15 and IL-6 were gathered immediately after dose two where these quantities have peaked, such that the subsequent decay was not characterized [19]. To ensure the eventual decrease in these cytokines as a function of time in our modelled prediction, we added an interval-censored data point on day 200, which ensures the fit on this day is between 0 and the minimal value determined on day 1 for each respective concentration.

## 3. Results

All standard two-dose BNT162b2 and mRNA-1273 IgG and cytokine data were simultaneously fit to our model (Eq. 1) using non-linear mixed-effects models in Monolix. The resulting population fit parameters are shown in Table S2, with all individual fit parameters shown in Tables S3 and S4. Plots of individual fits to each data set are shown in Figs S1-S3. In this work we also consider a comparison to two low doses (25 *μg*) of mRNA-1273. The population parameter fit values to the low dose data can be found in Table S2, while all individual fit values can be found in Tables S5-S6. Plots for individual fits to the two low doses of mRNA-1273 data can be found in Fig. S7.

### 3.1. Model is consistent with clinically-observed humoral responses

In Fig. 1 we summarize our findings for the IgG responses from two standard doses of BNT162b2 or mRNA-1273. To determine an average standard two-dose mRNA-1273 response from all of our individual fits we average the individual IgG two-dose mRNA-1273 fitted parameters from Tables S3 and S4 for Widge *et al*. [21] age groups 18-55, 56-70, 70+, and Wang *et al*. [22] RBD and Spike IgG. The result is shown as the green points with corresponding average fit in Fig. 1a. Similarly we do the same for the BNT162b2 two-dose result, where we average the individual IgG fitted parameters from Tables S3 and S4 from Stankov *et al*. [18], Bergamaschi *et al*. [19], Camara *et al*. [20], Wang *et al*. [22], and Suthar *et al*. [23]. The result is shown as the red points with corresponding average fit in Fig. 1a. The population fit (blue line) in Fig. 1a utilizes all data from Tables S3 and S4, corresponding to the population fitted parameters in Table S2. We show our long-term predicted response for each two-standard-dose vaccine type up to day 265, which marks the initial approximate date of the planned dose three, eight months following dose two [29], however, was later adjusted to a six month post-dose-two timeline [30].

**Figure 1:**
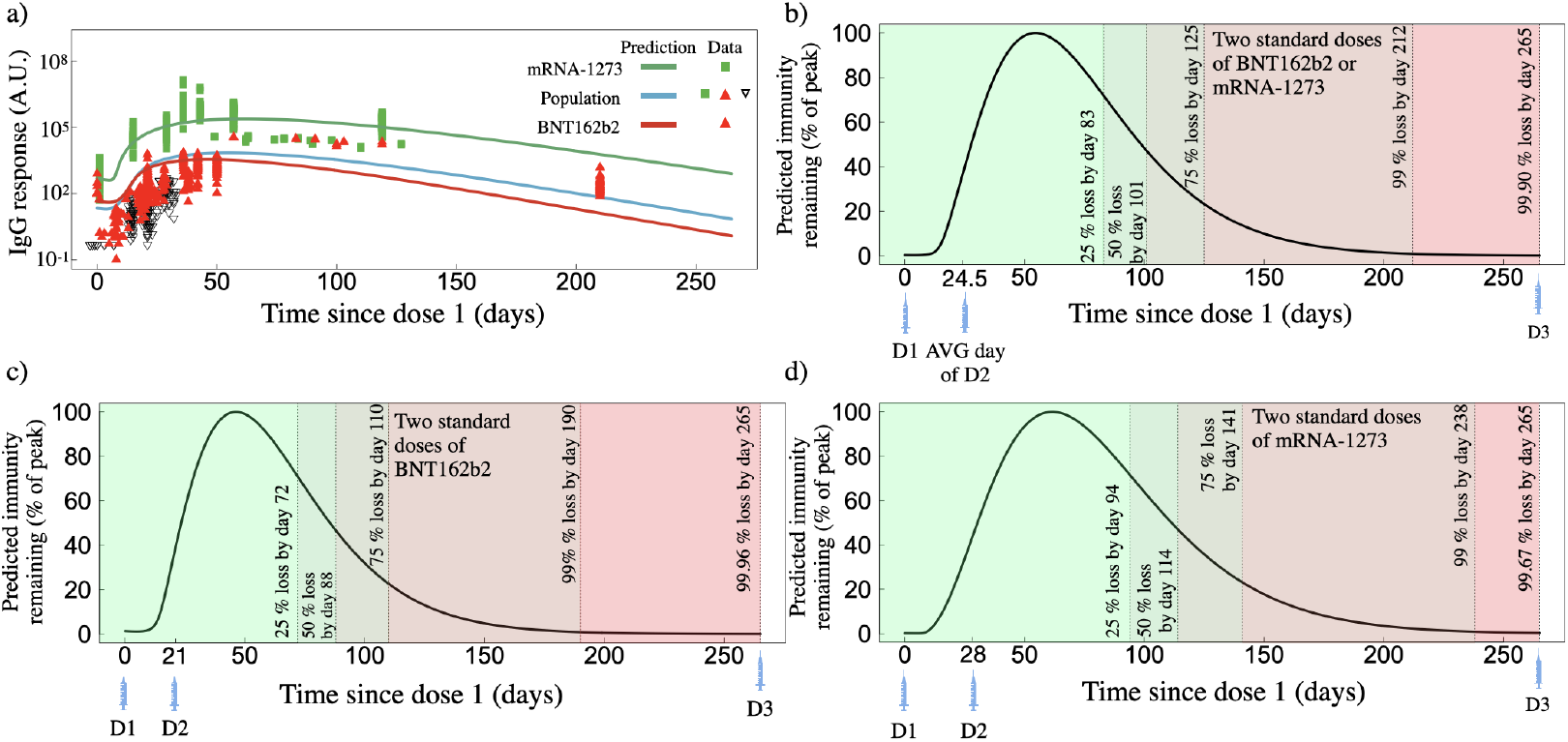
Predicted IgG response based on fits to clinical data for two standard doses of BNT162b2 or mRNA-1273. a) IgG as a function of time since dose one for two standard doses of mRNA-1273 (green line, green squares represent actual data), BNT162b2 (red line, red triangles represent actual data), and the population fit which uses both mRNA-1273 and BNT162b2 two-dose data (blue line, combines all available data). b) Predicted IgG remaining, normalized by the peak IgG count, for the population fit (panel a), blue line) as a function of days since the first dose. c) Predicted IgG remaining for the two-dose BNT162b2 fit (corresponding to red line in Fig. 1a) as a function of days since the first dose. d) Predicted IgG remaining for the two-dose mRNA-1273 fit (corresponding to green line in Fig. 1a) as a function of days since the first dose.

We note that for all IgG responses, the y-axes are in arbitrary units (A.U.) and depend on the methodology employed within each respective publication. In this work each clinical data set has not been re-normalized or adjusted. As such, direct comparison between the magnitude of the BNT162b2 or mRNA-1273 IgG responses can not be reasonably completed. However, the time dependence of the relaxation of each IgG response relative to their respective peak response can be directly compared. Each IgG fit is therefore normalized by its peak immunity determined through its respective fit. For Fig. 1b, c and d, the green to red colour sequence distinguishes 25, 50, 75, 99 and *>*99 IgG% loss relative to peak immunity. The time at which each percent loss occurs for each vaccine, as well as the humoral degradation rate, *γ*_*A*_, fit from Eq. 1e, are shown in Table 2.

**Table 2:**
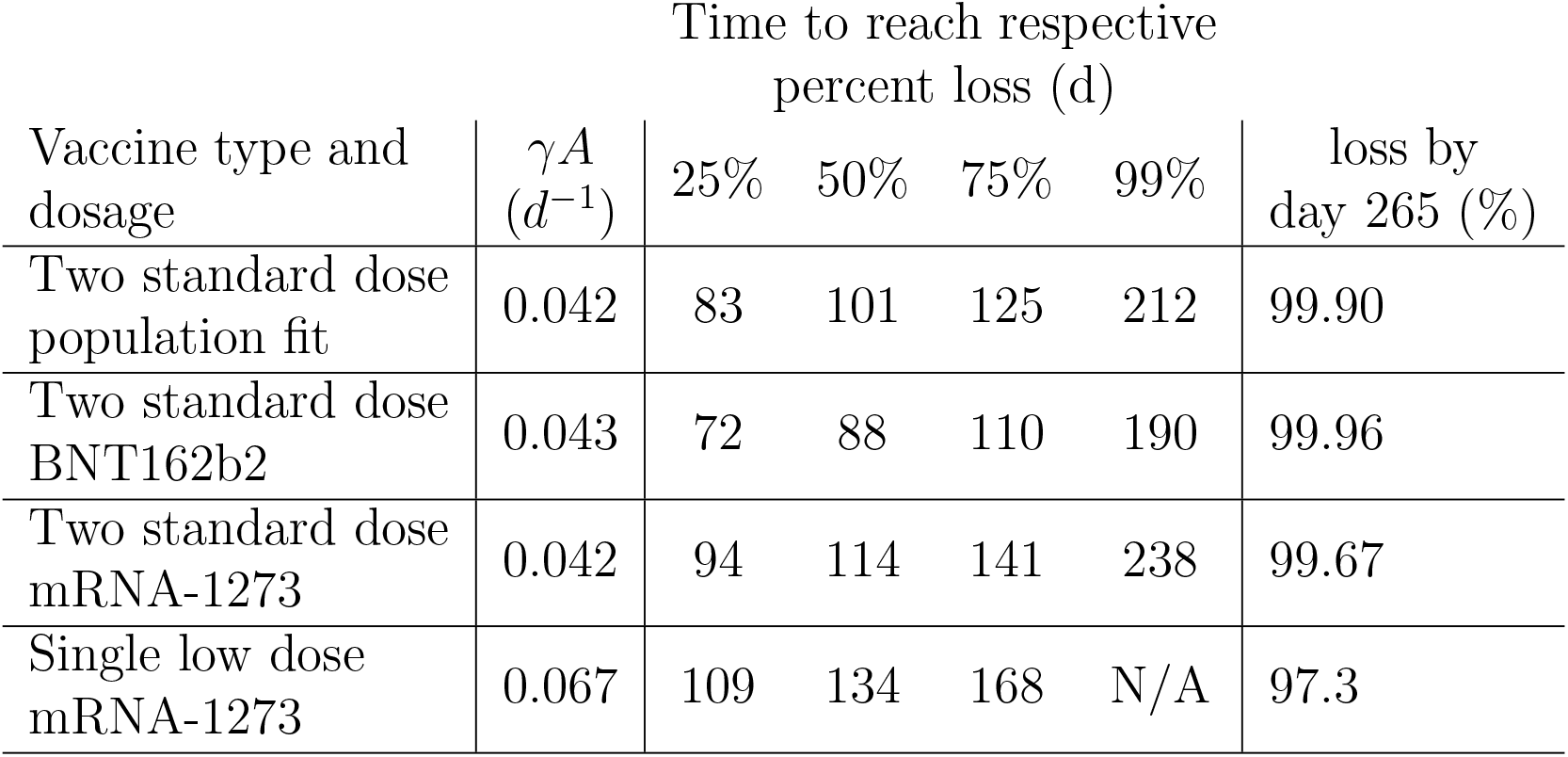
Summary of IgG predictions presented. Time to reach respective loss values are measured as time since the day of first dose.

We find that the humoral degradation rate *γ*_*A*_ for two standard doses of mRNA-1273 and BNT162b2 to be 0.043 and 0.042 *d*^−1^. The time to reach the 25, 50, 75, 99 and *>*99 percent loss milestones also varies substantially between vaccine types with 94, 114, 141, and 238 days for mRNA-1273, and 72, 88, 110 and 190 days for BNT162b2. By day 265 following dose one, we find both mRNA-1273 and BNT162b2 are predicted to lead to antibody counts less than 99.5% peak loss.

### 3.2. Long term predictions for two standard doses of mRNA-1273 depending on age

The clinical data used for our fit to two standard doses of mRNA-1273 includes a separation of their data by age [21]. Where as in Fig. 1a we plot the overall response from all age cohorts, in Fig. 2a we separate the age cohorts into 18-55, 55-70, and 70+ aged individuals and plot our model predicted responses. We find nearly identical long-term behaviour between the 56-70 and 70+ individuals with release rate *μ*_*BA*_ of 0.94 *d*^−1^ and 0.87 *d*^−1^, respectively, and *γ*_*A*_ of 0.045 *d*^−1^ and 0.045 *d*^−1^, respectively. For the 18-55 cohort we find *μ*_*BA*_ and *γ*_*A*_ to be 0.74 and 0.04 *d*^−1^, respectively. We also find differences in the plasma B cell death rates, *γ*_*B*_; younger individuals have a *γ*_*B*_ of 0.044 *d*^−1^ while 70+ individuals have a larger *γ*_*B*_ of 0.061 *d*^−1^ leading to faster plasma B cell degradation among older individuals. These age-dependent differences are leading to a higher IgG response in younger individuals by day 265.

**Figure 2:**
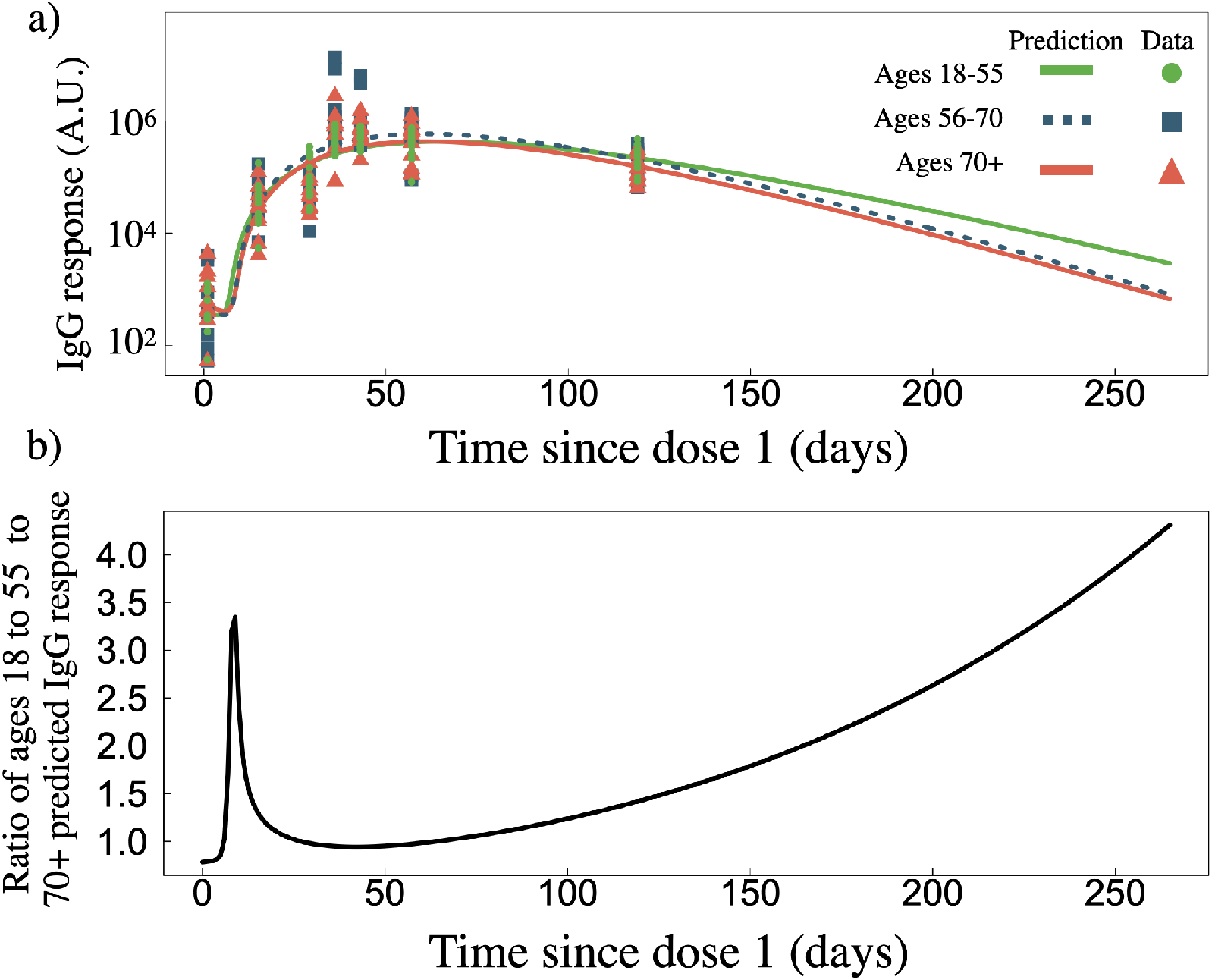
a) IgG response as a function of time since dose one for two standard doses of mRNA-1273, separated by age groups 18-55 (*n* = 15), 56-70 (*n* = 9), and 70+ (*n* = 10) years of age, with data sourced from ref. [21]. b) Ratio of predicted IgG response of 18-55 age cohort to the 70+ age cohort.

To better illustrate the differences between the predicted responses from 18-55 and 70+ individuals, we compared the ratio of 18-55 and 70+ fitted responses (Fig. 2b). We find that by day ∼50 the ratio of IgG response between 18-55 and 70+ age cohorts is ∼1, however, as time progresses an exponential advantage for the 18-55 cohort emerges, such that by day 265 the 18-55 age group has roughly four-fold more IgG than compared to the 70+ age group. We also find a much stronger initial IgG response in the 18-55 age cohort, where by day ten these individuals have 3 times more IgG, however, this advantage quickly decays. All fitted Eq. 1 model parameters for all age-specific predictions can be found Tables S3 and S4.

### 3.3. Sex dependant long term predictions for two standard doses of BNT162b2

Amongst the clinical data used for our fit to two standard doses of BNT162b2 is a data set where IgG response is separated by sex [23]. There are little differences amongst any fitted parameters between male and female individuals (Tables S3 and S4). In Fig. 3a we plot the sex-dependent predictions as well as the corresponding clinical data separated by sex. In Fig. 3b we plot the ratio of the male and female predicted response. A higher initial IgG advantage emerges for males that peaks on day 30, however, the immunity advantage slowly dissipates such that by day 265 the ratio of male to female IgG response is ∼1.

**Figure 3:**
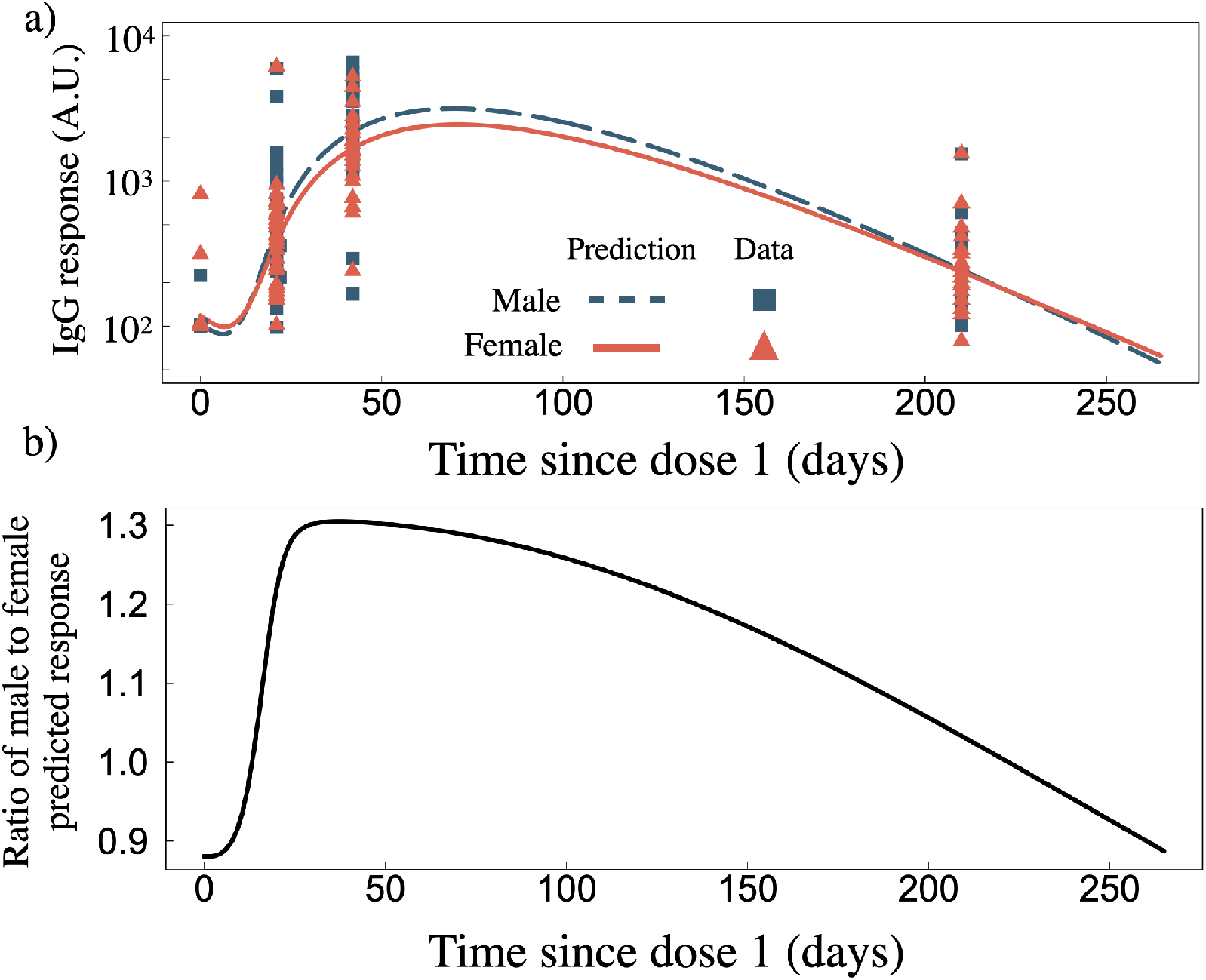
a) IgG response as a function of time since dose one for two standard doses of BNT162b2 for male (*n* = 22) and female (*n* = 24) data sets [23]. b) Ratio of male to female predicted IgG response from panel a).

### 3.4. Long term predictions for two low doses of mRNA-1273

Fig. 4a displays our IgG population model prediction for two low-doses of mRNA-127 (25*μ*g as opposed to 100*μ*g). The clinical data for these fits was sourced from ref. [24] (this data set was not included in the standard dose fits described above). Individual fits to the RBD and spike IgG can be found in Fig. S7, all individual fit parameters can be found in Tables S5 and S6, and our two-low-dose population fitted parameters can be found in Table S2 alongside the standard dose population fitted parameters. The population fit for two-low-doses mRNA-1273 yields *γ*_*A*_ = 0.067 *d*^−1^. The time to reach a percent loss of 25, 50, 75%, relative to peak IgG value is 109, 134, 168 days, and by day 265 we find 97.3% of the peak response has waned.

**Figure 4:**
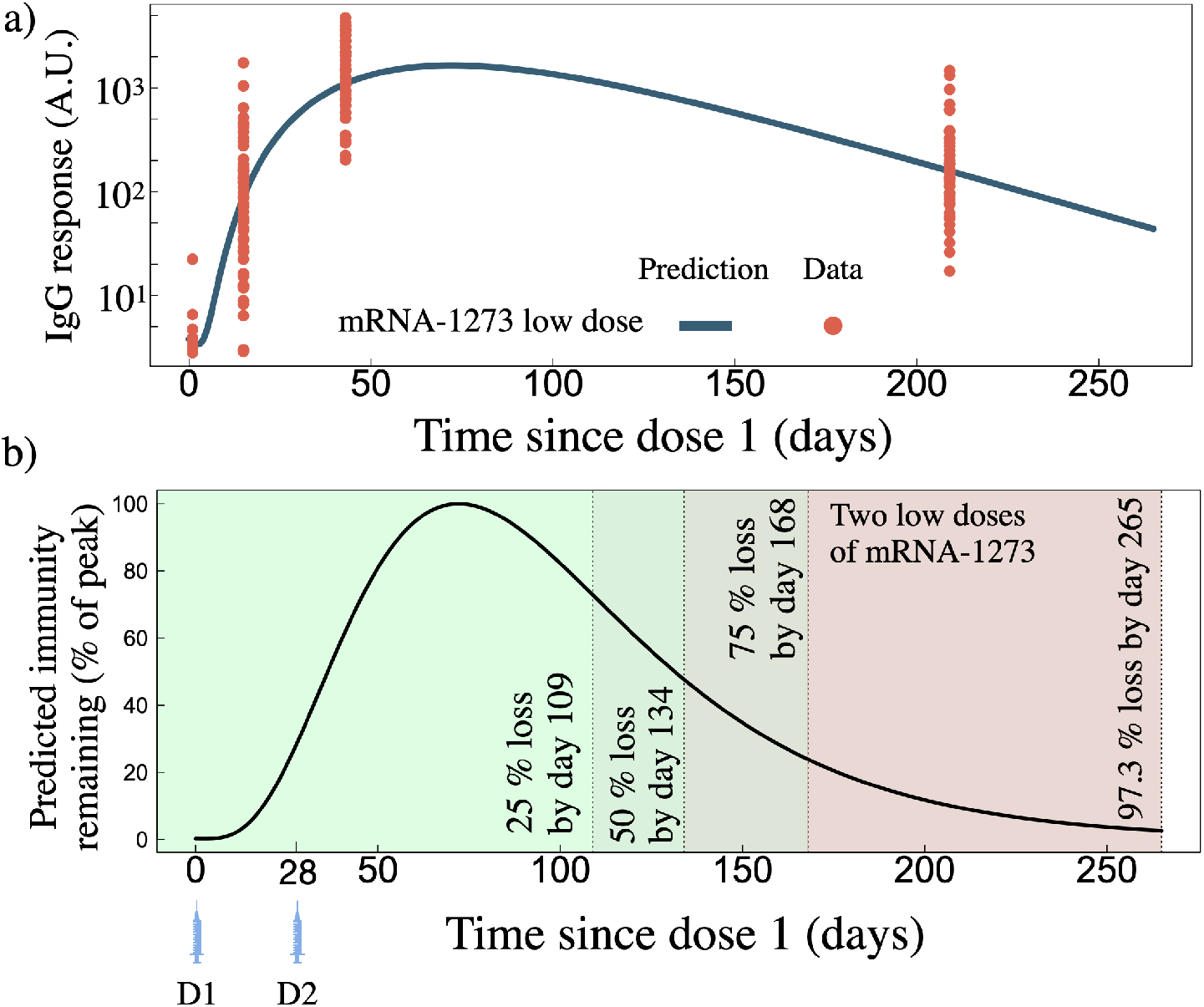
a) IgG as a function of time since dose one for two low doses of mRNA-1273. Blue line is the model prediction fit from the clinical data (*n* = 33) shown as orange points. b) Predicted IgG remaining, normalized by the peak IgG count, for the two low doses of mRNA-1273 shown in panel a).

## 4. Discussion

In this work, we have developed a novel within-host mathematical model (Eq. 1) to describe the vaccine dynamics from LNP-formulated mRNA vaccines. An in-depth description of our model is provided in the supplementary text. We then curated 22 previously published two-dose mRNA-1273 and BNT162b2 antibody and cytokine clinical results to fit to our model (Table 1). We simultaneously fit our model to two-standard doses of either mRNA-1273 or BNT162b2, and also separately fit our model to two-low doses of mRNA-1273. For all clinical data used in this work, the time between doses is the vaccine developer’s recommended dose interval time. Of the two-standard doses of mRNA-1273, we also perform our fits considering different age categories to examine how age affects the long-term predicted immune response where data is available to do so [21]. We further consider how sex affects the long-term BNT162b2 predictions where data was available to do so [23]. In line with the initial planned dose three interval set out by the CDC [29], eight months following dose two, we extend all of our fits to predict the response out to day 265 following dose one.

Table 2 summarizes our long-term predictions shown in Fig. 1a-d. We find humoral immunity gained from two standard doses of mRNA-1273 and BNT162b2 to peak on days 63 and 47 following dose one, respectively. We normalize our curves by the peak IgG value to then determine the milestones for percent loss relative to the peak. A number of modelling studies predicting waning immunity caution SARS-CoV-2 resurgence upon inadequate administration of a booster dose, depending on the severity of how quickly immunity wanes [31, 32, 33]. Before dose three is administered, we predict a significant window of time where individuals will have less than 99% humoral loss relative to peak immunity of 105 and 27 days for BNT162b2 and mRNA-1273, respectively. Our predictions are therefore in agreement with the CDC’s adjusted timeline of six months post dose two for dose three [30], as opposed to the initial eight month post dose two plan [29]. Overall, we find that two doses of mRNA-1273 to decay slower relative to peak immunity compared to two doses of BNT162b2 (Table 2).

This modest humoral advantage as a function of time for mRNA-1273 recipients is in line with the recent CDC MMWR report which found vaccine effectiveness in preventing hospitalization was higher for Moderna recipients compared to Pfizer [34]. Furthermore, individuals who received two-doses of mRNA-1273 have been found to have dramatically reduced neutralizing antibody activity six months following dose two [35]; suggesting a six month timeline for dose three after dose two [30].

An immunologic correlate of protection against SARS-CoV-2 has not yet been established [36]. Our model, however, allows us to make a clinically-guided qualitative prediction for a humoral-based immunologic correlate of protection. For example, it has been found that seven months following dose two of Pfizer/Biontech efficacy in preventing infection drops from 75% to 16% [10]. At seven months past dose two for BNT162b2 we predict IgG levels to drop to 0.16 percent of the peak response; suggesting that 0.16% of the peak response correlates to 16% efficacy. In another BNT162b2 study, efficacy against infection was found to decline to 47% five months post dose two, with efficacy against the delta variant found to be 53% four months after full vaccination [37]. We find that following two standard doses of BNT162b2 the IgG counts have dropped to 2.1 and 7.2 percent of peak, five and four months following the second dose, respectively.

An antibody study of mRNA-1273 found vaccine efficacy as low as 50.8%, and as high as 96.1%, 28 days past dose two, and further found vaccine efficacy to increase throughout day 29 to 57 past dose one [38]. We find that 28 days past dose two mRNA-1273 IgG levels are predicted to be at 99% of their peak value, and from days 29 to 57 past dose one the IgG levels are predicted to increase towards the peak value, which occurs on day 63 (Fig. 1d).

It is known that as individuals age, they can develop numerous possible molecular immunological impairments that lead to an inability to maintain humoral immunity [39, 40]. Therefore, we expanded our study to predict humoral immune loss as a function of age for two-standard-dose mRNA-1273. We find that over the first 20 days post dose one the younger cohort has a much stronger predicted IgG response compared to the 70+ cohort, such that by day 10 they have achieved a three-fold IgG advantage over the older cohort (Fig. 2b). A more rapid antibody response to vaccination is not unexpected in younger individuals, where factors such as impaired B cell development and maintenance are more common in the elderly [41]. Indeed, we find that plasma B cells are produced at roughly the same rate amongst all ages, however, die at a slower rate of 0.044 d^−1^ in younger individuals, compared to a rate of 0.061 d^−1^ in older individuals; which suggests that younger individuals are better maintaining their plasma B cell population.

The initial humoral advantage in younger individuals quickly dissipates, such that by day 50 the ratio of IgG response between 18-55 and 70+ individuals is ∼ 1.0. As time progresses beyond day 50 after dose one, we find there exists an increased ability to maintain humoral immunity amongst 18-55 aged individuals compared to 56-70 and 70+ aged individuals (Fig. 2b), such that by day 265 the 18-55 aged individuals are predicted to have ∼ 4-fold more IgG than compared to older individuals. This result is in line with mRNA-1273 low-dose clinical findings where a two-fold reduction in IgG by day 209 in older cohorts was found [24]. These results support a dose three roll out strategy to prioritize older individuals first, as we predict their humoral immune loss to occur faster compared to younger individuals.

As the world’s nations respond to the rapidly spreading SARS-CoV-2 pandemic and the emergence of its many variants, vaccine conservation is becoming increasingly important. As cautioned by the World Health Organization, many individuals in some priority populations have not yet received a primary vaccination [42]. This begs the question; will a lower dose elicit protection against a SARS-CoV-2 challenge? Without an immunologic correlate of protection against SARS-CoV-2 this question is difficult to answer explicitly. However, towards addressing the concern, we fit our model to two-low-dose (25*μ*g) mRNA-1273 data sets, and compared the degradation kinetics relative to peak immunity to our two-standard-dose results. All population fit parameters for standard and low dose scenarios are shown side-by-side in Table S2. For the low dose fits we find antibody degradation to be approximately ∼ 1.6 times faster, while the released antibody rate is a factor of ∼ 1.1 times slower, as compared to the standard dose population fits. We find, however, that relative to peak immunity, the two-low-dose strategy loses immunity slower, such that by day 265 since dose one recipients are predicted to have 97.3% loss of peak immunity, compared to *>* 99% peak loss for two standard doses of mRNA-1273 or BNT162b2 (Table 2).

## 5. Summary

In our study we develop a novel within-host mathematical model to describe the vaccination process of LNP-formulated mRNA vaccines, and we fit our model to 22 mRNA-1273 and BNT162b2 clinical data sets to determine best-fit model parameters and establish accurate long term predictions. We separate our individual fits by age, sex, and vaccine type. We find young individuals (18-55 of age) to be more responsive to vaccination as well as maintain humoral immunity longer than compared to older (70+ of age) individuals. Our results suggest males have a slightly higher peak response to two doses of BNT162b2, however, there exists little difference between sexs in the ability to maintain humoral immunity over the long term. We predict that two standard doses of either vaccine results in less than 99% of peak immunity remaining by day 190 and 238 past dose one for BNT162b2 and mRNA-1273, respectively. Relative to peak IgG response, the the mRNA-1273 vaccine is found to decay slower as a function of time as compared to BNT162b2.

Our results will help guide public health policies regarding booster dose three timelines. Our humoral results, correlated with efficacy against infection studies, are in agreement with the CDC timeline that a booster of an LNP-formulated mRNA vaccine may be required within 6-8 months of the second dose to maintain high effectiveness against SARS-CoV-2 and the emerging variants [30, 29].

## Data Availability

All data produced in the present study are available upon reasonable request to the authors.

## 6. Contributors

JMH, HKO and CSK conceived the project and designed the study. CSK completed the statistical analysis and fitting in Monolix as well as model predictions in **R**. All authors had the opportunity to discuss the results and comment on the manuscript. All authors had full access to all the data in the study and had final responsibility for the decision to submit for publication.

## 8. Data Sharing

All data that support the findings of this study are available within the manuscript and the supplementary information files or from the corresponding authors upon reasonable request.

## 9. Acknowledgements

We thank the Canadian In-host Modelling group (Matt Betti, Dan Coombs, David Dick, Thomas Hillen, Jude Kong, Kang Ling Liao, Nicole Mideo, Stéephanie Portet, Angie Raad, Lindi Wahl, James Watmough) for fruitful discussions.

