## Supplemental Material for "Long-term predictions of humoral immunity after two doses of BNT162b2 and mRNA-1273 vaccines based on dosage, age and sex"

October 13, 2021

Chapin S. Korosec, Suzan Farhang Sardroodi, David W. Dick, Samaneh Gholami, Mohammad Sajjad Ghaemi, Iain R. Moyles, Morgan Craig, Hsu Kiang Ooi, and Jane M. Heffernan

Corresponding author emails:

### 1 Supporting Text

#### 1.1 Model for in-host mRNA vaccination

Here we describe our model for mRNA vaccination delivered by lipid nano particles, and the subsequent in-host immunization process. We model the time dependence of eight state variables: lipid nanoparticles ($L$), vaccinated cells ($V$), $CD4^+$ cells ($T$), plasma B cells ($B$), antibody ($A$), $CD8^+$ cells ($C$), and the cytokines interferon ($F$) and interleukin ($I$). The model developed in this work is adapted from our recently published adenovirus-based model [1].

##### 1.1.1 Inoculation and vaccination

BNT162b2 and mRNA-1273 both contain base-modified — or nucleoside-modified — *bm*RNA that encode for full-length diproline-stabilized SARS-COV-2 viral spike protein, and are delivered via a payload enclosed by a lipid nanoparticle (LNP) [2,3]. The standard mRNA dose in BNT162b2 is 30$\mu$g, and together with the known mRNA size of 4.3 kb [2] and average nucleotide molecular weight of 319 $g \cdot mol$, there are an estimated 1.32x$10^{13}$ of $bm$RNA in each dose. The standard dosage of mRNA-1273 contains 100$\mu$g of $bm$RNA, while the molecular weight of the molecule is not public knowledge. LNP-based mRNA theraputics are a novel techonology where the pathway to activating the innate and and adpative immune response in humans is not well understood. Further, the route of administration of an LNP-based vaccine has a pronouced effect on the targeted cell types and tissues; for example, a luciferase-based mouse study by Pardi *et al.* found that intradermally, intraperitoneally, subcutaneously, intramuscularly, and intravenously inocculated LNP vaccines have widely varying outcomes [4]. While intraperitoneally and intravenously delivered LNP vaccines tended to primarily drain to hepatocytes in the liver, intramuscularly, subcutaneously, and intradermally delivered LNP vaccine stayed near the site of injection for many days (with partial drainage to the liver). A later study examining intramuscular injection in mice found that an LNP-based mRNA vaccine activated the innate immune system and remained active near the injection site as well as in draining lymph nodes [5]. The cytokines TNF and IL-6 were found in the muscle as as well as in the draining lymph nodes, as well as activation of B cells, and CD8+, CD4+ T cells. Motivated by such studies, the SARS-CoV-2 mRNA-based vaccines BNT162b2 or mRNA-1273 are delivered intramuscularly to humans.

Upon receiving a dosage of BNT162b2 or mRNA-1273, the patient is therefore likely producing spike protein near the site of injection for many days, as well as in lymph nodes for a period of time. The timescales and activation thresholds between the innate and adaptive immune response, driven by the proportions of the LNP dose absorbed into myocytes, draining lymph nodes, and hepatocytes is not well known. Therefore, in this work, we take a course grained approach to the in-host immunization process and model a general 'vaccinated' cell compartment.

The mechanistic model we propose for innoculation and preceding immunization in-host from an LNP-based mRNA vaccine is as follows. The number of LNPs in-host after inoculation is

$$\frac{d\mathrm{L}}{dt} = -\mu_{L,V}\mathrm{L} - \gamma_{\mathrm{L}}\mathrm{L}. \tag{1}$$

The $\mu_{L,V}$ term accounts for the interaction between the LNP-mRNA payload and target cell, $V$. We consider a cell successfully vaccinated upon the successful fusion of a LNP, leading to subsequent expression of the spike protein (which is not explicitly modelled). $\gamma_{\mathrm{L}}\mathrm{L}$ captures the natural degradation of LNPs in-host, as well as fusion inefficiency and drainage of the LNPs to the liver. The vaccinated cell count is then

$$\frac{d\mathrm{V}}{dt} = \mu_{L,V}\mathrm{L} - \gamma_V V, \tag{2}$$

where $\gamma_V$ is the natural death rate of the vaccinated cells.

##### 1.1.2 CD4$^+$ cell priming by vaccinated cells

Having received the mRNA payload vaccinated cells express antigen, leading to the development of membrane-bound class-II peptide-major histocompatibility complexes (MHC-II). Naive CD4$^+$ T-helper cells (given by $T$ in our model) recognize and bond with MHC-II where an information exchange occurs [6]. These dynamics are captured by the $\mu_{T,V}$ term in our model. The CD4$^+$ cell is then considered primed with antigen information. We therefore model the activation of naive CD4$^+$ cells by

$$\frac{dT}{dt} = \mu_{T,V}V - \gamma_T T, \tag{3}$$

where $\mu_{T,V}$ is the rate of interaction between naive $T$ cells and vaccinated cells and $\gamma_T$ is the natural death rate of $T$ cells. In this work $\mu_{T,V}$ is determined through a fit to clinical data. The death rate is fixed to 0.055 d$^{-1}$ as determined from by a clinical study [7].

##### 1.1.3 Humoral immunity

We model a primary antibody response generated by plasma B cells ($B$). Plasma B cells are primed by antigen-specific T cells [8], and in our model is taken into account by the interaction rate $\mu_{TB}$. The maturation of a naive B cell into an antibody secreting cells is further enhanced by various cytokines such as interleukin [8,9]. We incorporate interleukin-mediated plasma B cell maturation into our model through the term $\alpha_{_{BI}}\left(\frac{I}{S_I+I}\right)$. $\alpha_{_{BI}}$ regulates Plasma B cell stimulation by interleukin, while $S_{_I}$ provides a duplication threshold of plasma B cells due to interleukin. We fix $s_{_I} = 1000$, which was determined by the two-dose half-max interleukin threshold in our previous work on adenovirus vaccines [1], and is further justified as accurate for this current work by the maximum clinical IL-8 values determined in ref. [10]. We therefore model the production of antigen-specific plasma B cells, $B$, as

$$\frac{dB}{dt} = \mu_{T,B}T + \alpha_{_{BI}}\left(\frac{I}{s_{_I}+I}\right)B - \gamma_B B, \tag{4}$$

where $\gamma_B$ is the natural death rate of plasma $B$ cells. The production of interleukin (I) by CD4$^+$ cells is described in section 1.1.4.

In this work we focus on the production of the predominant antibody in humans immunoglobulin G (IgG) [11], and do not distinguish between subclasses of IgG or consider other types of immunoglobulin. In our model IgG is given by $A$, and we model the production of $A$ by $B$ through the source term $\mu_{B,A}B$. The rate of change of $A$ is then

$$\frac{dA}{dt} = \mu_{_{BA}}B - \gamma_A A, \tag{5}$$

where $\gamma_A A$ is the natural antibody degradation term.

##### 1.1.4 CD8$^+$ cell priming and cytokine response

CD8$^+$ Cyotoxic T cells, denoted by $C$ in our model, target and eliminate virus-infected cells. CD8$^+$ cells are primed by antigen presenting cells (Eq. 2) through cell-surface bound MHC class I molecules [12]. In our model this process is captured by the source term $\mu_{CV}V$. The regulation of antigen-specific CD8$^+$ by cytokines is complex. Type I IFN has been shown to enhance the CD8+ T cell response during priming [13], and Type II IFN (IFN-$\gamma$) has been directly shown to enhance development of CD8+ memory [14, 15]. We therefore model the enhancement of CD8+ memory development through a source term dependent on the presence of IFN-$\gamma$ (denoted by $F$ in our model).

The rate of change of of CD8$^+$, $C$, cells is then

$$\frac{dC}{dt} = \mu_{CV}V + \alpha_{C,F}\left(\frac{F}{S_F + F}\right)C - \gamma_C C, \tag{6}$$

where $\gamma_C$ is the natural death rate of CD8$^+$ cells, $\alpha_{C,F}$ accounts for stimulation by $F$, and $S_F$ is the duplication threshold due to $F$. In this work we fix $s_F = 600$ which was determined by the two-dose half-max IFN$-\gamma$ threshold in our previous work on adenovirus vaccines [1], and is further justified as accurate based on the two-dose IFN-$\gamma$ data used in this work from refs. [16] and [10].

CD4$^+$ and CD8$^+$ cells exhibit complex cytokine secretion and regulation dynamics. Here, we consider a simplified approach where IFN$-\gamma$ and Interleukin ($I$) cytokine production is accomplished by primed CD4$^+$ cells (eq. 3). T helper cells are one the predominant sources of cytokine production [17]; CD4+ cells have been shown to secrete IFN-$\gamma$ [18,19] and are one of the primary synthesizers of interleukins [20]. We therefore model the production of IFN-$\gamma$ ($F$) and interleukin ($I$) by

$$\frac{dF}{dt} = \mu_{TF}T - \alpha_{FC}CF - \gamma_F F, \tag{7}$$

and

$$\frac{dI}{dt} = \mu_{TI}T - \alpha_{IB}IB - \gamma_I I. \tag{8}$$

The natural degradation of $F$ and $I$ is described by $\gamma_F$, and $\gamma_I$, respectively. Upon CD8+ enhancement by $F$ we then model the subsequent clearance of $F$ by $\alpha_{FC}CF$. Similarly, upon Plasma B cell enhance by $I$ we model the subsequent clearance of $I$ by $\alpha_{IB}IB$; thus we do not allow $F$ and $I$ to enhance the development of multiple cells.

The complete model we use to describe the in-host immunization process by LNP-formulated mRNA vaccines is then given by

$$\frac{d\mathrm{L}}{dt} = -\mu_{_{LV}}\mathrm{L} - \gamma_{\mathrm{L}}\mathrm{L} \tag{9a}$$

$$\frac{d\mathrm{V}}{dt} = \mu_{_{LV}}\mathrm{L} - \gamma_{_V}V \tag{9b}$$

$$\frac{dT}{dt} = \mu_{_{TV}}V - \gamma_{_T}T \tag{9c}$$

$$\frac{dB}{dt} = \mu_{_{TB}}T + \alpha_{_{BI}}\left(\frac{I}{s_I + I}\right)B - \gamma_{_B}B \tag{9d}$$

$$\frac{dA}{dt} = \mu_{_{BA}}B - \gamma_{_A}A \tag{9e}$$

$$\frac{dC}{dt} = \mu_{_{CV}}V + \alpha_{_{CF}}\left(\frac{F}{s_F + F}\right)C - \gamma_{_C}C \tag{9f}$$

$$\frac{dF}{dt} = \mu_{_{TF}}T - \alpha_{_{FC}}CF - \gamma_{_F}F \tag{9g}$$

$$\frac{dI}{dt} = \mu_{_{TI}}T - \alpha_{_{IB}}IB - \gamma_{_I}I. \tag{9h}$$

Table 1.1.4 summarizes the state variable definitions, units and initial conditions.

| Variable | Definition | Units | Initial condition |
|---|---|---|---|
| $L$ | Lipid nano particle | A.U. | 1 |
| $V$ | Vaccinated cell | A.U. | 0 |
| $T$ | CD4+ T cell | A.U. | 0 |
| $B$ | Plasma B cell | A.U. | 0 |
| $A$ | Antibodies | A.U | $A_0$ |
| $C$ | CD8+ T cell | A.U. | 0 |
| $F$ | IFN-$\gamma$ | pg/ml | 0 |
| $I$ | Interleukin | pg/ml | $I_0$ |

Table S1: Model variables and initial conditions for individual and population fits used throughout this work. We simultaneously fit to multiple IgG data sets, where the data sets come from different labs and have arbitrary units (a.u.). We therefore fit an initial condition A given by $A_0$. We furthermore simultaneously fit to 4 different interleukin data sets, each having varying responses through time and initial dynamics. We therefore fit the initial condition $I_0$. The initial conditions in this table represent the *population* initial condition from our fits; every individually-fitted data set may have slightly varying initial dynamics.

#### 2 All population fit values for two standard doses and two low doses

The population fit values are determined through a fit to all individual data data sets use for each vaccine dosage regimen. Values and figures for every individual fit for each data set are shown in the following sections.

| | | Population fit values | | |
|---|---|---|---|---|
| Parameter | Definition | Two standard doses | Single low dose | Comment |
| $\mu_{LV}$ | LNP absorption rate with antigen presenting cells | 0.91 | 0.023 | Fit |
| $\gamma_L$ | LNP degradation rate | 0.00013 | 0.00026 | Fit |
| $\gamma_V$ | Antigen presenting cell death rate | 0.07 | 0.098 | Fit |
| $\mu_{TV}$ | CD4+ activation rate by vaccinated cells | 4.98 | 3.86 | Fit |
| $\gamma_T$ | CD4+ natural death rate | 0.055 | 0.055 | Ref. [7] |
| $\mu_{TB}$ | Plasma B cell activation rate by CD4+ cells | 0.098 | 2.48 | Fit |
| $\alpha_{BI}$ | Plasma B cell stimulation by Interleukin | 2.4 | 0.019 | Fit |
| $S_I$ | Plasma B cell duplication threshold due to Interleukin | 1000 | 1000 | Chosen |
| $\gamma_B$ | Plasma B cell natural death rate | 0.071 | 0.072 | Fit |
| $\mu_{BA}$ | Released antibody rate by plasma B cells | 0.51 | 0.48 | Fit |
| $\gamma_A$ | Antibody natural degradation rate | 0.042 | 0.067 | Fit |
| $\mu_{CV}$ | CD8+ activation rate by vaccinated cells | 0.000022 | 0.00033 | Fit |
| $\alpha_{CF}$ | CD8+ stimulation by IFN-$\gamma$ | 0.0000014 | 0.00096 | Fit |
| $S_F$ | CD8+ duplication threshold due to IFN-$\gamma$ | 600 | 600 | Chosen |
| $\gamma_C$ | CD8+ natural death rate | 0.01 | 0.01 | Ref. [21] |
| $\mu_{TF}$ | IFN-$\gamma$ stimulation rate by Thelper cells | 194.79 | 0.22 | Fit |
| $\alpha_{FC}$ | IFN-$\gamma$ clearance by cytotoxic Tcells | 0.0000013 | 0.000004 | Fit |
| $\gamma_F$ | IFN-$\gamma$ natural degradation rate | 201.24 | 64.82 | Fit |
| $\mu_{TI}$ | Interleukin secretion by CD4+ cells | 2.07 | 0.000057 | Fit |
| $\alpha_{IB}$ | Interleukin clearance by Plasma B cells | 0.0019 | 1.07 | Fit |
| $\gamma_I$ | Natural interleukin degradation rate | 0.027 | 0.00001 | Fit |
| $A_0$ | Antibody initial condition | 22.5 | 3.8 | Fit |
| $I_0$ | Interleukin initial condition | 5.18 | 1.0 | Fit |
| BIC | Bayesian Information Criteria | 3468 | 315 | Fit |
| AIC | Akaike Information Criteria | 3421 | 252 | Fit |

Table S2: Model parameters definition and population fitted values for two standard doses of BNT162b2 or mRNA-1273 and two low doses of mRNA-1273. The dosing times are separated by 21 and 28 days for BNT162b2 and mRNA-1273, respectively.

### 3 Two standard doses mRNA vaccination of BNT162b2 or mRNA-1273

All individual fits shown in this section were fit simultaneously in monolix.

#### 3.1 Individual data set fitted values

| Data set ID | Figure reference, quantity used | Vaccine | $\mu_{LV}$ | $\gamma_L$ | $\gamma_V$ | $\mu_{TV}$ | $\gamma_T$ | $\mu_{TB}$ | $\alpha_{BI}$ | $S_I$ | $\gamma_B$ | $\mu_{BA}$ | $\gamma_A$ |
|---|---|---|---|---|---|---|---|---|---|---|---|---|---|
| Goel at al. [22] | Fig. 1b. RBD IgG | BNT162b2 & mRNA-1273 | 0.77 | 0.00012 | 0.07 | 0.75 | 0.054 | 0.07 | 2.67 | 1000 | 0.18 | 0.27 | 0.042 |
| Goel et al. [22] | Fig. 1b, Spike IgG | BNT162b2 & mRNA-1273 | 1.08 | 0.00012 | 0.07 | 1.34 | 0.055 | 0.077 | 2.42 | 1000 | 0.12 | 0.5 | 0.043 |
| Stankov et al. [23] | Fig. 1a, Spike IgG | BNT162b2 | 0.86 | 0.0001 | 0.081 | 9.42 | 0.068 | 0.092 | 2.84 | 1000 | 0.31 | 0.07 | 0.048 |
| Bergamaschi et al. [10] | Fig. 1a, Spike-RBD IgG, | BNT162b2 | 1.08 | 0.00013 | 0.079 | 1.97 | 0.065 | 0.078 | 2.32 | 1000 | 0.21 | 0.91 | 0.047 |
| Camara et al. [16] | Fig. 1b, Spike IgG, | BNT162b2 | 0.94 | 0.00013 | 0.083 | 4.05 | 0.067 | 0.082 | 2.63 | 1000 | 0.21 | 0.22 | 0.048 |
| Bergamaschi et al. [10] | Fig. 2A, IFN-$\gamma$ | BNT162b2 | 3.88 | 0.00012 | 0.067 | 4.02 | 0.051 | 0.098 | 2.41 | 1000 | 0.071 | 0.51 | 0.042 |
| Camara et al. [16] | Fig. 1a, IFN-$\gamma$, | BNT162b2 | 1.14 | 0.00012 | 0.057 | 8.21 | 0.044 | 0.098 | 2.39 | 1000 | 0.071 | 0.5 | 0.042 |
| Bergamaschi et al. [10] | Fig. 2c, IL-6 | BNT162b2 | 0.36 | 0.00011 | 0.071 | 0.81 | 0.059 | 0.094 | 2.39 | 1000 | 0.073 | 0.53 | 0.042 |
| Bergamaschi et al. [10] | Fig. 2b, IL-8 | BNT162b2 | 1.04 | 0.00012 | 0.13 | 7.16 | 0.11 | 0.088 | 4.44 | 1000 | 0.026 | 0.45 | 0.042 |
| Bergamaschi et al. [10] | Fig. 2a, IL-15 | BNT162b2 | 0.45 | 0.00012 | 0.068 | 0.78 | 0.054 | 0.096 | 2.39 | 1000 | 0.074 | 0.53 | 0.042 |
| Widge et al. [24] | Fig. 1a (RBD antibody, 18-55 yrs) | mRNA-1273 | 0.9 | 0.00012 | 0.06 | 12.44 | 0.048 | 0.096 | 2.43 | 1000 | 0.044 | 0.74 | 0.04 |
| Widge et al. [24] | Fig. 1a (RBD antibody, 56-70 yrs) | mRNA-1273 | 0.17 | 0.00011 | 0.066 | 14.55 | 0.057 | 0.083 | 2.13 | 1000 | 0.062 | 0.94 | 0.045 |
| Widge et al. [24] | Fig. 1a (RBD antibody, 70+ yrs) | mRNA-1273 | 0.17 | 0.00013 | 0.066 | 12.73 | 0.056 | 0.1 | 2.25 | 1000 | 0.061 | 0.87 | 0.045 |
| Bergamaschi et al. [10] | Fig. 2b (IL-16) | BNT162b2 | 1.64 | 0.00012 | 0.067 | 26.84 | 0.053 | 0.11 | 2.2 | 1000 | 0.54 | 0.54 | 0.042 |
| Wang et al. [25] | Fig. 1e (RBD IgG) | mRNA-1273 | 0.97 | 0.00012 | 0.064 | 6.36 | 0.049 | 0.098 | 2.45 | 1000 | 0.044 | 0.56 | 0.039 |
| Wang et al. [25] | Fig. 1f (Spike IgG) | mRNA-1273 | 0.95 | 0.00012 | 0.064 | 6.57 | 0.048 | 0.099 | 2.45 | 1000 | 0.043 | 0.57 | 0.039 |
| Wang et al. [25] | Fig. 1e (RBD IgG) | BNT162b2 | 0.93 | 0.00012 | 0.066 | 7.25 | 0.052 | 0.099 | 2.47 | 1000 | 0.057 | 0.59 | 0.04 |
| Wang et al. [25] | Fig. 1f (Spike IgG) | BNT162b2 | 1.05 | 0.00012 | 0.065 | 7.2 | 0.05 | 0.098 | 2.47 | 1000 | 0.053 | 0.6 | 0.04 |
| Suthar et al. [26] | Fig. 1a (Spike IgG, Male) | BNT162b2 | 1.2 | 0.00012 | 0.068 | 6.57 | 0.051 | 0.11 | 2.36 | 1000 | 0.03 | 0.38 | 0.039 |
| Suthar et al. [26] | Fig. 1a (Spike IgG, Female) | BNT162b2 | 0.99 | 0.00012 | 0.072 | 5.23 | 0.054 | 0.13 | 2.2 | 1000 | 0.026 | 0.42 | 0.039 |

Table S3: Individual fit values for to the various data sets used in this work. This table contains all fitted parameters from equations 9a, 9b, 9c, 9d, and 9e.

| Data set ID | Paper Fig. reference, quantity used | Vaccine | $\mu_{CV}$ | $\alpha_{CF}$ | $S_F$ | $\gamma_C$ | $\mu_{TF}$ | $\alpha_{FC}$ | $\gamma_F$ | $\mu_{TI}$ | $\alpha_{IB}$ | $\gamma_I$ | $A_0$ | $I_0$ |
|---|---|---|---|---|---|---|---|---|---|---|---|---|---|---|
| Goel at al. [22] | Fig. 1b, RBD IgG | BNT162b2 & mRNA-1273 | 0.000021 | 1.4E-06 | 600 | 0.0078 | 194.83 | 1.3E-06 | 202.39 | 2.97 | 0.0019 | 0.022 | 0.58 | 2.14 |
| Goel et al. [22] | Fig. 1b, Spike IgG | BNT162b2 & mRNA-1273 | 0.000022 | 1.4E-06 | 600 | 0.014 | 195.27 | 1.3E-06 | 199.53 | 1.14 | 0.0016 | 0.024 | 0.52 | 3.74 |
| Stankov et al. [23] | Fig. 1a, Spike IgG | BNT162b2 | 0.000024 | 1.4E-06 | 600 | 0.018 | 185.85 | 1.3E-06 | 196.98 | 1.33 | 0.0035 | 0.026 | 1.12 | 3.23 |
| Bergamaschi et al. [10] | Fig. 1a, Spike-RBD IgG | BNT162b2 | 0.00002 | 1.4E-06 | 600 | 0.0083 | 193.89 | 1.3E-06 | 207.77 | 2.34 | 0.0013 | 0.024 | 3.69 | 3.54 |
| Camara et al. [16] | Fig. 1b, Spike IgG | BNT162b2 | 0.000022 | 1.4E-06 | 600 | 0.0028 | 195.21 | 1.3E-06 | 202.22 | 1.35 | 0.0023 | 0.026 | 2.88 | 3.52 |
| Bergamaschi et al. [10] | Fig. 2A, IFN-$\gamma$ | BNT162b2 | 0.000022 | 1.4E-06 | 600 | 0.0094 | 189.29 | 1.3E-06 | 266.97 | 2.27 | 0.0019 | 0.027 | 22.85 | 5.34 |
| Camara et al. [16] | Fig. 1a, IFN-$\gamma$ | BNT162b2 | 0.000022 | 1.4E-06 | 600 | 0.01 | 207.17 | 1.3E-06 | 124.78 | 2.07 | 0.0019 | 0.027 | 23.4 | 4.96 |
| Bergamaschi et al. [10] | Fig. 2c, IL-6 | BNT162b2 | 0.000024 | 1.4E-06 | 600 | 0.0093 | 191.21 | 1.3E-06 | 230.1 | 0.035 | 0.0018 | 0.05 | 18.39 | 0.89 |
| Bergamaschi et al. [10] | Fig. 2b, IL-8 | BNT162b2 | 0.00002 | 1.4E-06 | 600 | 0.011 | 193.87 | 1.3E-06 | 212.17 | 12.28 | 0.0017 | 0.027 | 26.26 | 0.65 |
| Bergamaschi et al. [10] | Fig. 2a, IL-15 | BNT162b2 | 0.000022 | 1.4E-06 | 600 | 0.011 | 192.95 | 1.3E-06 | 200.71 | 0.049 | 0.0018 | 0.035 | 23.43 | 2.06 |
| Widge et al. [24] | Fig. 1a (RBD antibody, 18-55 yrs) | mRNA-1273 | 0.000022 | 1.4E-06 | 600 | 0.01 | 195.09 | 1.3E-06 | 200.28 | 15.79 | 0.0016 | 0.028 | 404.79 | 4.95 |
| Widge et al. [24] | Fig. 1a (RBD antibody, 56-70 yrs) | mRNA-1273 | 0.000021 | 1.4E-06 | 600 | 0.004 | 194.37 | 1.3E-06 | 224.13 | 25.77 | 0.0014 | 0.028 | 440.37 | 5.18 |
| Widge et al. [24] | Fig. 1a (RBD antibody, 70+ yrs) | mRNA-1273 | 0.000026 | 1.4E-06 | 600 | 0.02 | 192.49 | 1.3E-06 | 195.36 | 22.97 | 0.0015 | 0.028 | 515.75 | 3.44 |
| Bergamaschi et al. [10] | Fig. 2b (IL-16) | BNT162b2 | 0.000022 | 1.4E-06 | 600 | 0.0089 | 195.54 | 1.3E-06 | 203.12 | 13.22 | 0.0021 | 0.027 | 21.99 | 235.56 |
| Wang et al. [25] | Fig. 1e (RBD IgG) | mRNA-1273 | 0.000022 | 1.4E-06 | 600 | 0.009 | 194.74 | 1.3E-06 | 201.76 | 3.52 | 0.0018 | 0.027 | 50.9 | 5.21 |
| Wang et al. [25] | Fig. 1f (Spike IgG) | mRNA-1273 | 0.000022 | 1.4E-06 | 600 | 0.01 | 195 | 1.3E-06 | 200.52 | 3.45 | 0.0018 | 0.027 | 50.82 | 5.13 |
| Wang et al. [25] | Fig. 1e (RBD IgG) | BNT162b2 | 0.000022 | 1.4E-06 | 600 | 0.01 | 194.79 | 1.3E-06 | 201.13 | 4.09 | 0.0018 | 0.027 | 50.87 | 5.16 |
| Wang et al. [25] | Fig. 1f (spike IgG) | BNT162b2 | 0.000021 | 1.4E-06 | 600 | 0.01 | 195.24 | 1.3E-06 | 199.48 | 4.1 | 0.0018 | 0.027 | 51.08 | 5.33 |
| Suthar et al. [26] | Fig. 1a (Spike IgG, Male) | BNT162b2 | 0.000021 | 1.4E-06 | 600 | 0.012 | 195.2 | 1.3E-06 | 200.65 | 0.43 | 0.0022 | 0.028 | 104.2 | 6.21 |
| Suthar et al. [26] | Fig. 1a (Spike IgG, Female) | BNT162b2 | 0.000022 | 1.4E-06 | 600 | 0.012 | 195.21 | 1.3E-06 | 201.71 | 0.43 | 0.0025 | 0.028 | 118.33 | 7.31 |

Table S4: Individual fit values for to the various data sets used in this work. This table contains all fitted parameters from equations 9f, 9g and 9h.

#### 3.2 Individual fits to IgG data sets

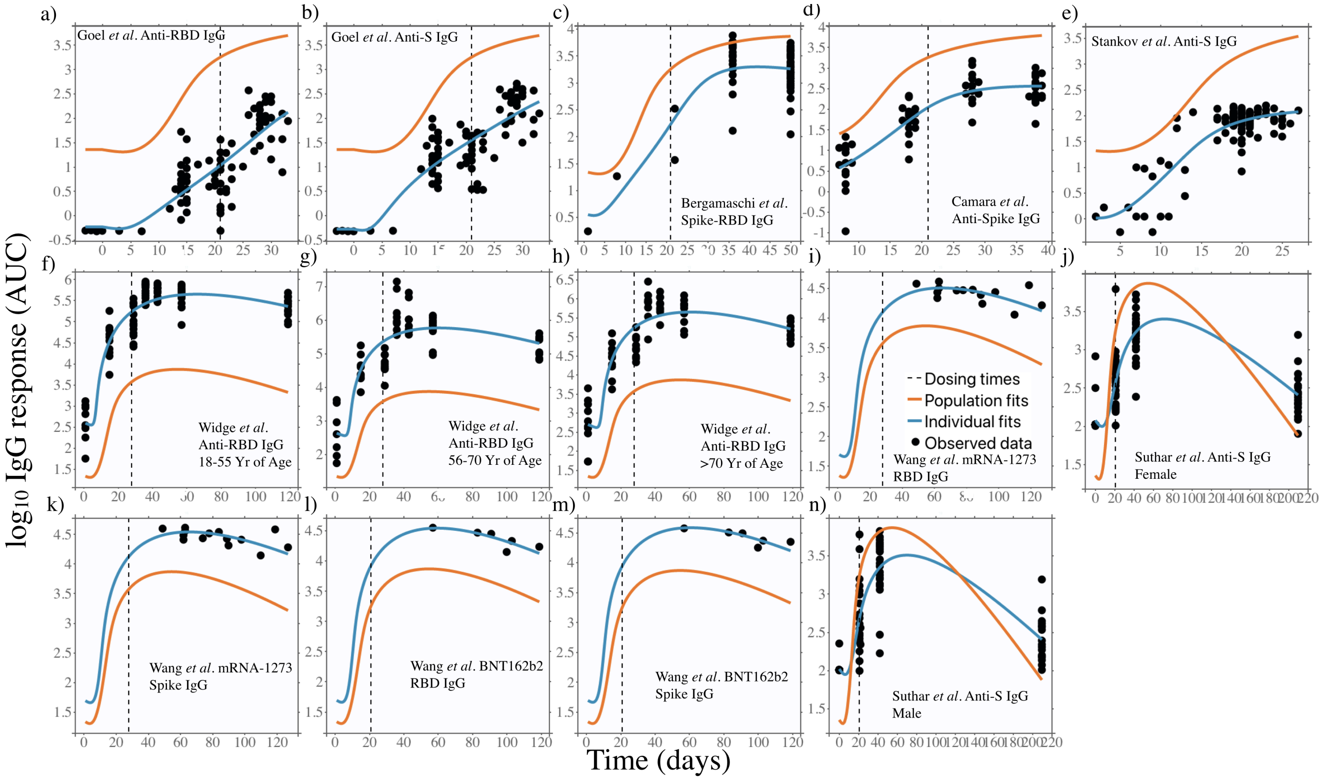

Figure S1: Individual fits to all standard dose IgG data sets used in this work. References for the data set sources can be found in Table 1 of the main text, all individual fitted parameters for each fit can be found in Tables S3 and S4.

##### 3.3 Individual fits to IFN-$\gamma$ data sets

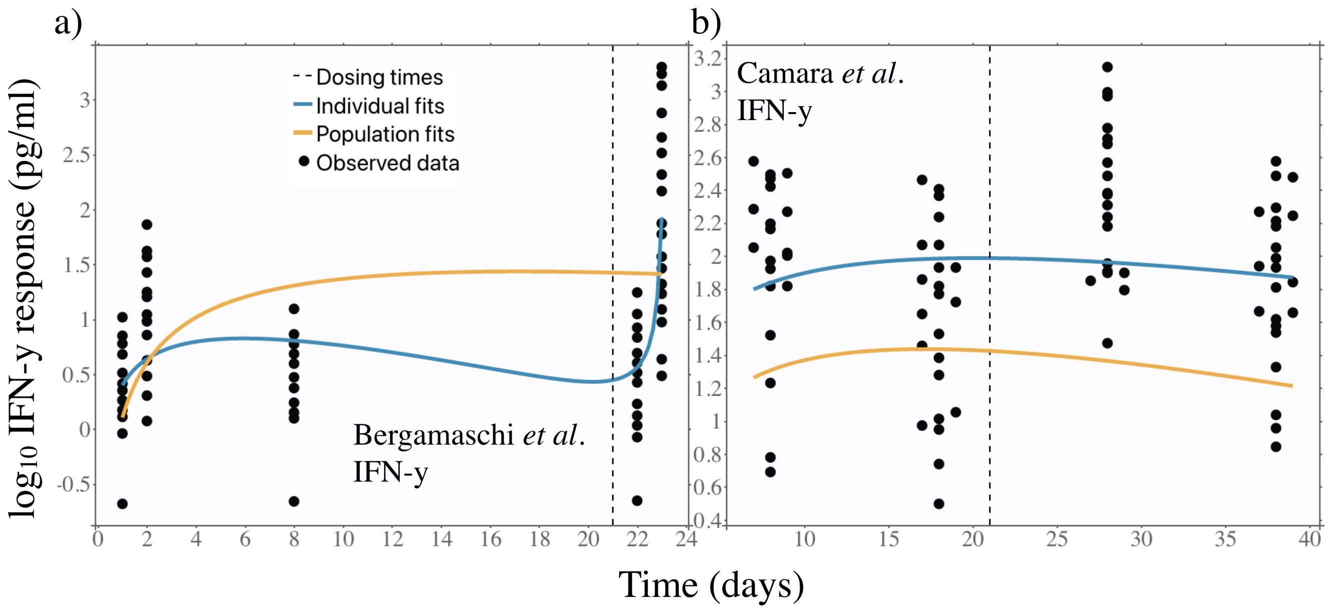

Figure S2: Individual fits to various IFN-$\gamma$ data sets. References for the data set sources can be found in Table 1 of the main text, all individual fitted parameters for each fit can be found in Tables S3 and S4.

##### 3.4 Individual fits to Interleukin data sets

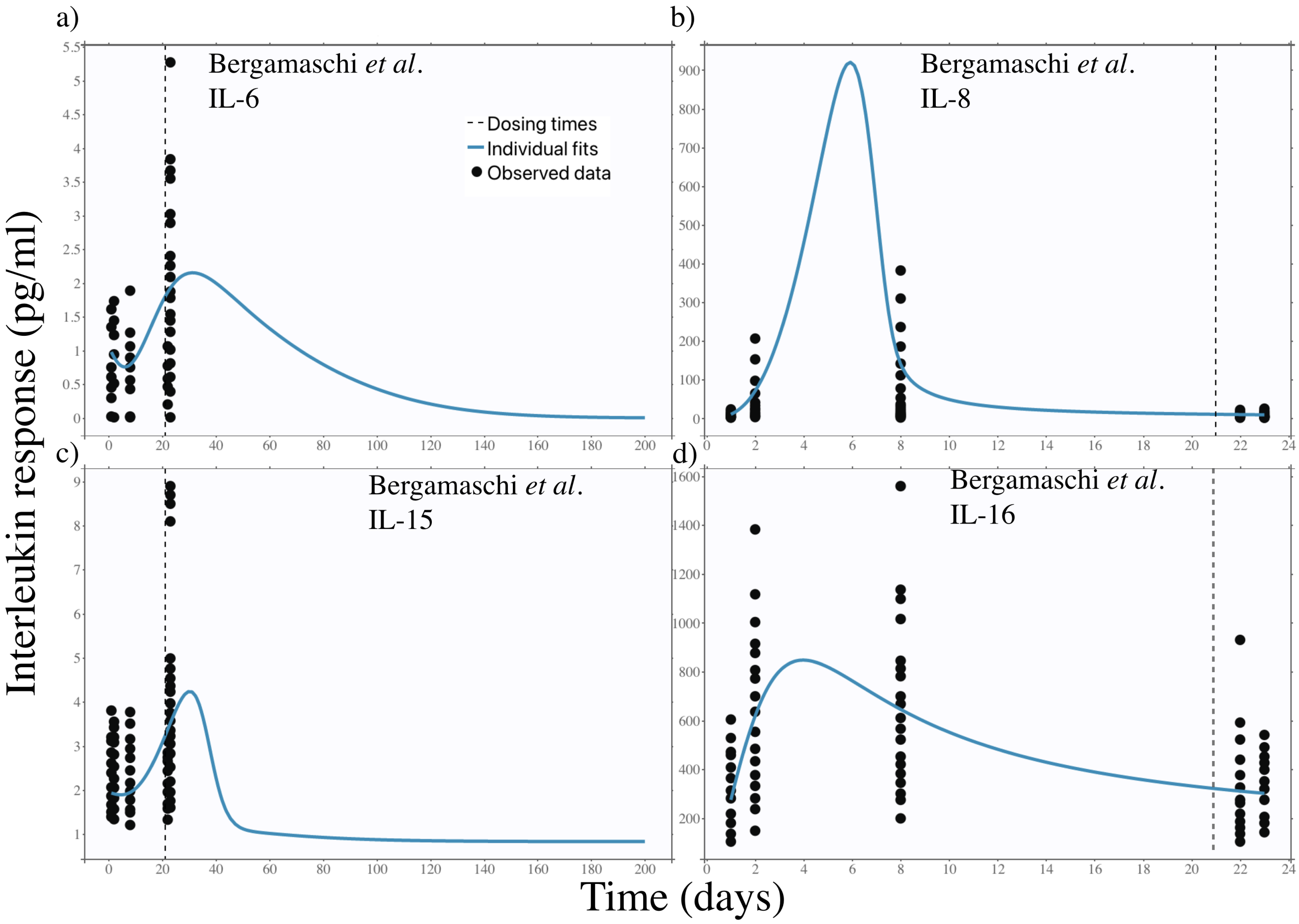

Figure S3: Individual fits to various interleukin data sets. References for the data set sources can be found in Table 1 of the main text, all individual fitted parameters for each fit can be found in Tables S3 and S4.

##### 3.5 Goodness of fit predictive checks and parameter distributions

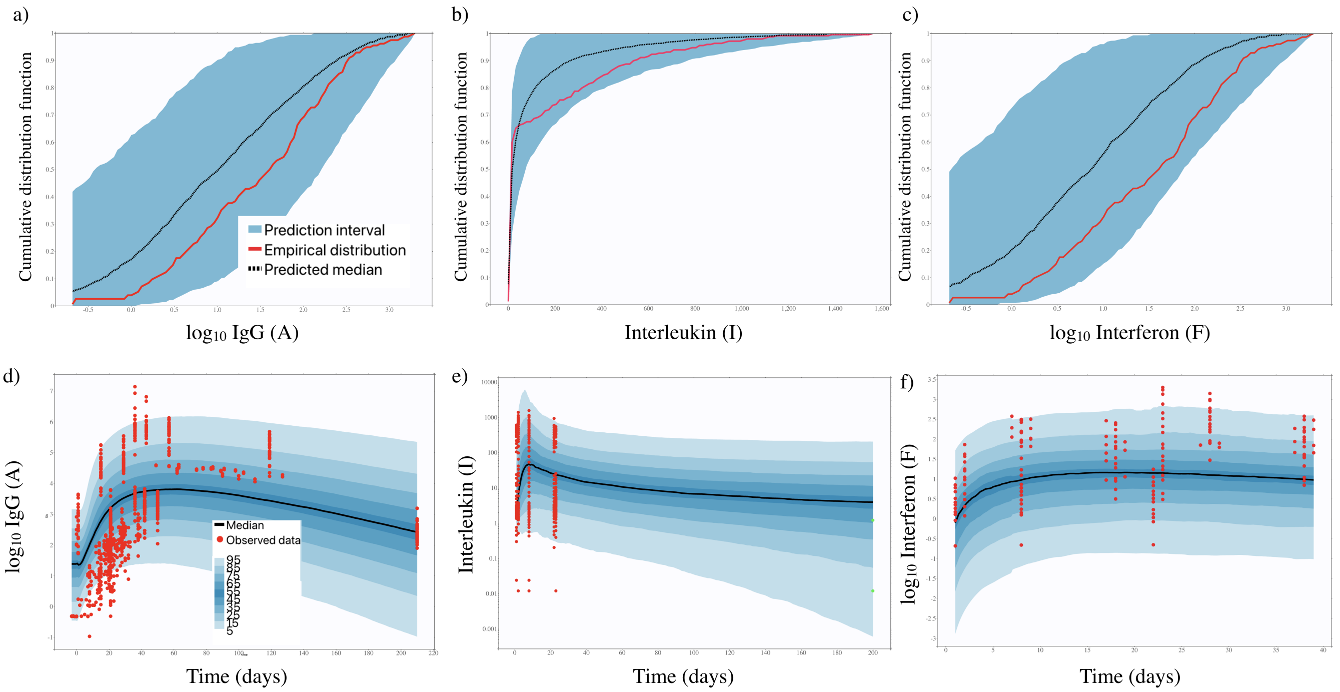

Figure S4: Predictive checks for two-dose mRNA results. a)-c) Numerical predictive checks for IgG, Interleukin, and Interferon, corresponding to fits to equations 9e, 9g, and 9h, respectively. d)-f) Prediction distributions for IgG, Interleukin, and Interferon as a function of time.

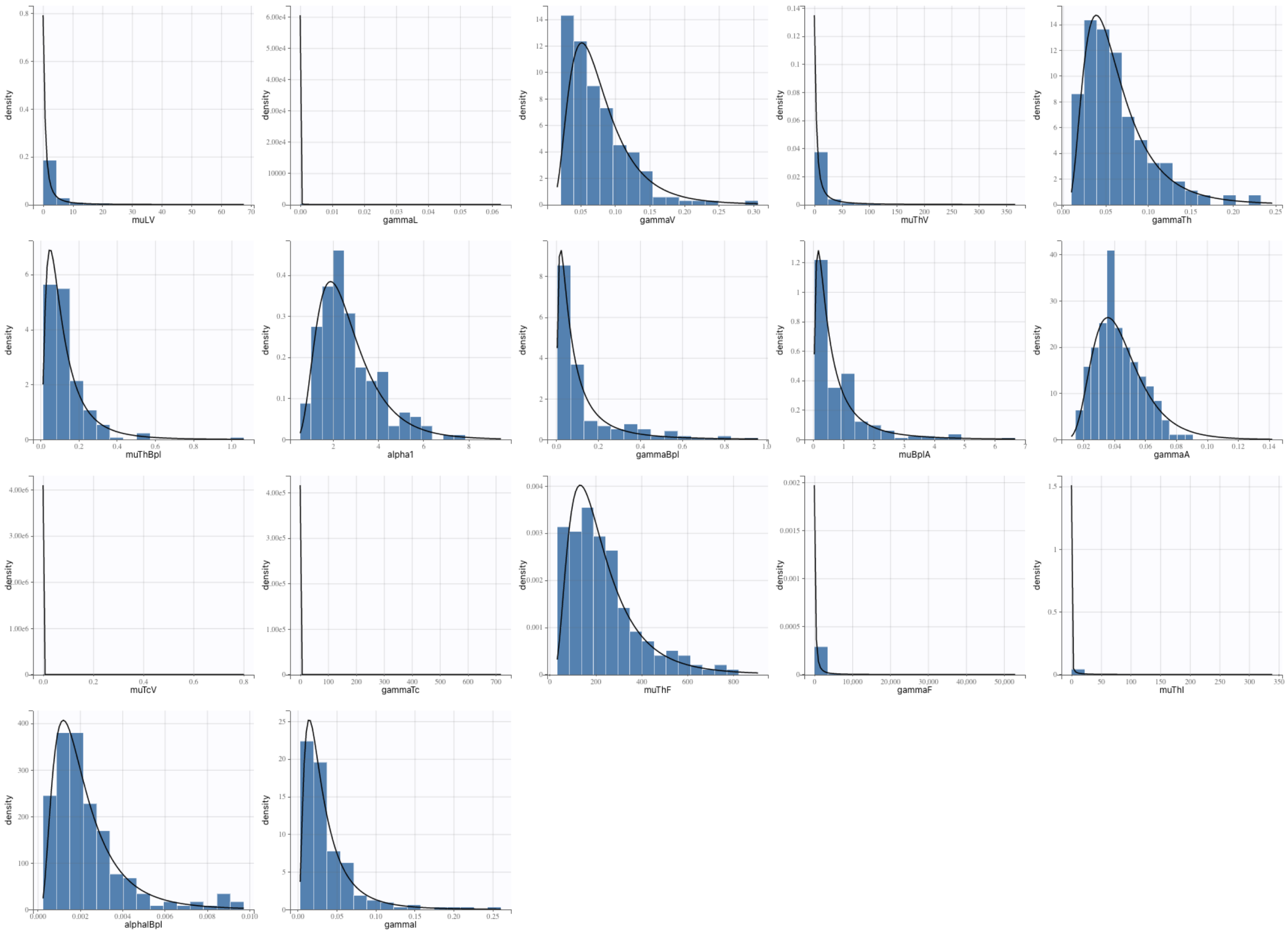

Figure S5: Monolix-determined fitted parameter distributions for all two standard dose fitted parameters.

### 4 Two low doses of mRNA-1273 vaccination

Two-low-dose mRNA-1273 data used in this work is sourced from Ref. [27]. This section contains tables summarizing the individual data set fitted parameters.

##### 4.1 Model parameter population fits and individual data set fitted values

| Data set ID | Figure from paper | $\mu_{LV}$ | $\gamma_L$ | $\gamma_V$ | $\mu_{TV}$ | $\gamma_T$ | $\mu_{TB}$ | $\alpha_{BI}$ | $s_I$ | $\gamma_B$ | $\mu_{BA}$ | $\gamma_A$ |
|---|---|---|---|---|---|---|---|---|---|---|---|---|
| Mateus *et al.* [27] | Fig. 1a (Spike IgG) | 0.023 | 0.00027 | 0.099 | 3.85 | 0.056 | 2.48 | 0.019 | 1003.37 | 0.073 | 0.48 | 0.068 |
| Mateus *et al.* [27] | Fig. 1b (RBD IgG) | 0.023 | 0.00025 | 0.097 | 3.87 | 0.054 | 2.49 | 0.019 | 1002.3 | 0.071 | 0.48 | 0.066 |

Table S5: Individual fit values for to the two low doses of mRNA-1273. This table contains all fitted parameters from equations 9a, 9b, 9c, 9d, and 9e.

| Data set ID | $\mu_{CV}$ | $\alpha_{CF}$ | $S_F$ | $\gamma_C$ | $\mu_{TF}$ | $\alpha_{FC}$ | $\gamma_F$ | $\mu_{TI}$ | $\alpha_{IB}$ | $\gamma_I$ |
|---|---|---|---|---|---|---|---|---|---|---|
| Mateus *et al.* [27] | 0.00033 | 0.00096 | 608.69 | 0.0099 | 0.21 | 0.000004 | 64.94 | 0.000057 | 1.07 | 0.0000074 |
| Mateus *et al.* [27] | 0.00033 | 0.00096 | 604.89 | 0.0099 | 0.2 | 0.000004 | 64.94 | 0.000056 | 1.07 | 0.0000074 |

Table S6: Individual fit values for to the two low doses of mRNA-1273. This table contains all fitted parameters from equations 9f, 9g and 9h.

##### 4.2 Individual fits to IgG data sets

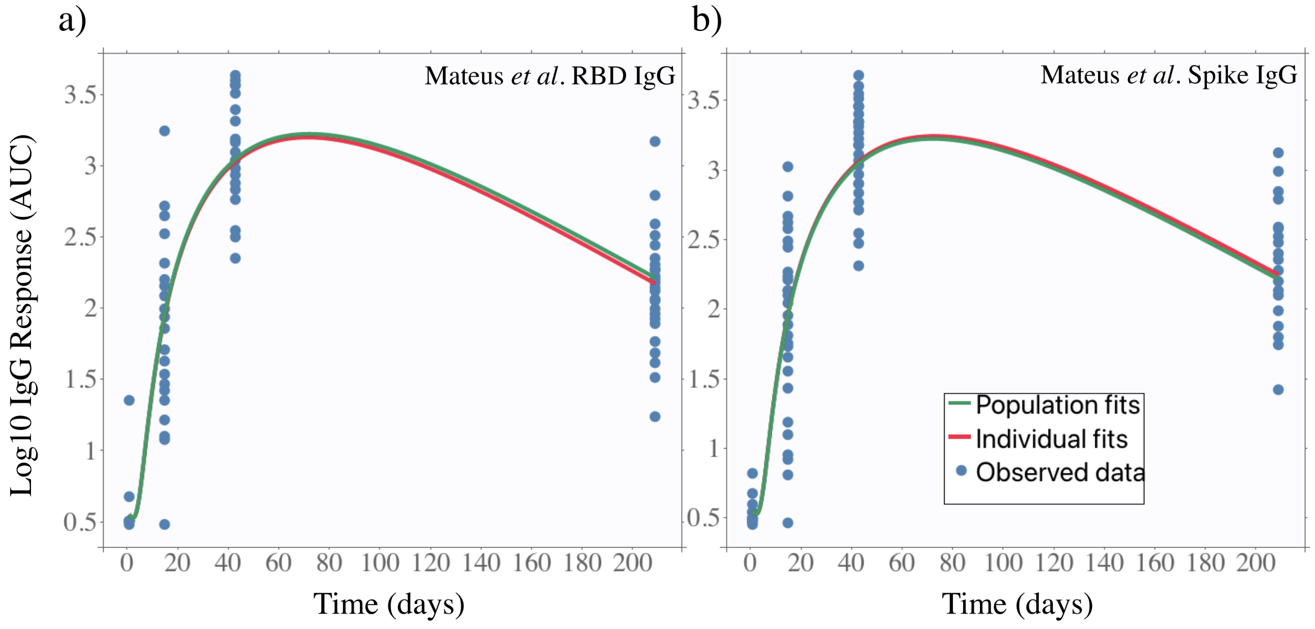

Figure S6: Individual fits to the two low doses of mRNA-1273. All individual fitted parameter values can be found in Tables S5 and S6.

#### 4.3 Goodness of fit predictive checks

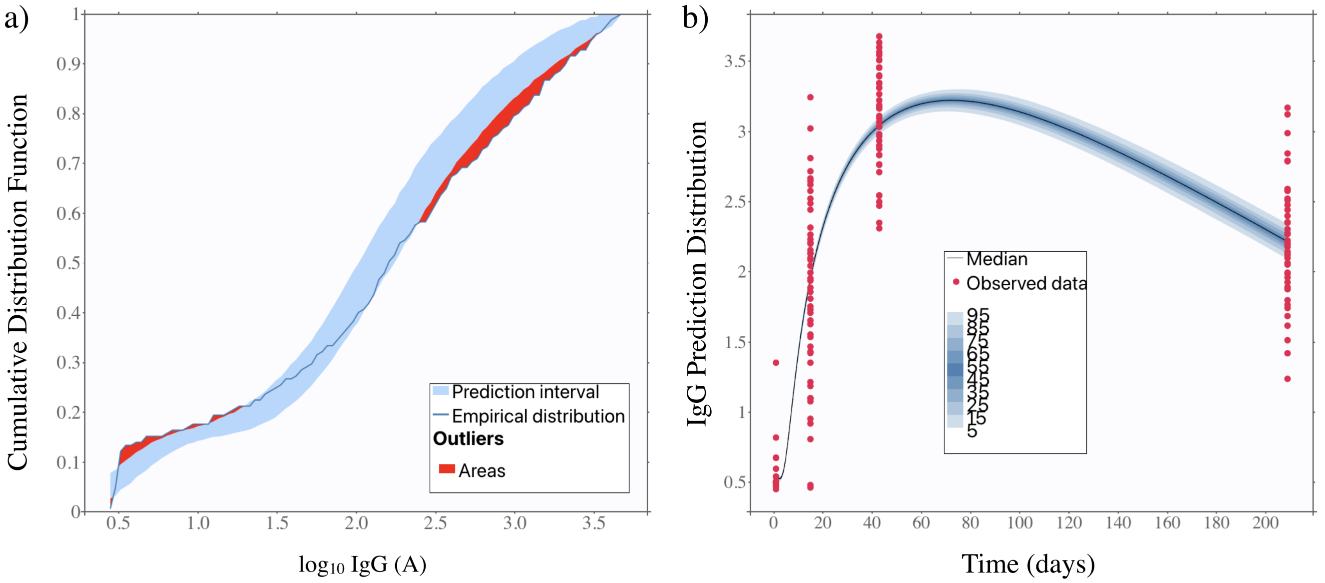

Figure S7: Predictive checks for fits to two low doses of mRNA-1273. a) Numerical predictive checks for the IgG data sets. b) Prediction distributions for the IgG fits as a function of time.

[3] H. Steinle, A. Behring, C. Schlensak, H. P. Wendel, M. Avci-Adali, S. F. Ahmed, A. A. Quadeer, M. R. McKay, N. N. Zhang, X. F. Li, Y. Q. Deng, H. Zhao, Y. J. Huang, G. Yang, W. J. Huang, P. Gao, C. Zhou, R. R. Zhang, Y. Guo, S. H. Sun, H. Fan, S. L. Zu, Q. Chen, Q. He, T. S. Cao, X. Y. Huang, H. Y. Qiu, J. H. Nie, Y. Jiang, H. Y. Yan, Q. Ye, X. Zhong, X. L. Xue, Z. Y. Zha, D. Zhou, X. Yang, Y. C. Wang, B. Ying, C. F. Qin, A. B. Vogel, I. Kanevsky, Y. Che, K. A. Swanson, A. Muik, M. Vormehr, L. M. Kranz, K. C. Walzer, S. Hein, A. Güler, J. Loschko, M. S. Maddur, K. Tompkins, J. Cole, B. G. Lui, T. Ziegenhals, A. Plaschke, D. Eisel, S. C. Dany, S. Fesser, S. Erbar, F. Bates, D. Schneider, B. Jesionek, B. Sänger, A.-K. Wallisch, Y. Feuchter, H. Junginger, S. A. Krumm, A. P. Heinen, P. Adams-Quack, J. Schlereth, C. Kröner, S. Hall-Ursone, K. Brasky, M. C. Griffor, S. Han, J. A. Lees, E. H. Mashalidis, P. V. Sahasrabudhe, C. Y. Tan, D. Pavliakova, G. Singh, C. Fontes-Garfias, M. Pride, I. L. Scully, T. Ciolino, J. Obregon, M. Gazi, R. Carrion, K. J. Alfson, W. V. Kalina, D. Kaushal, P.-Y. Shi, T. Klamp, C. Rosenbaum, A. N. Kuhn, Ö. Türeci, P. R. Dormitzer, K. U. Jansen, U. Sahin, W. Tai, L. He, X. Zhang, J. Pu, D. Voronin, S. Jiang, Y. Zhou, L. Du, T. Schlake, A. Thess, M. Fotin-Mleczek, K. J. Kallen, N. Pardi, M. J. Hogan, F. W. Porter, D. Weissman, S. Xu, K. Yang, R. Li, L. Zhang, S. Guan, J. Rosenecker, I. ModernaTX, P. S. Kowalski, A. Rudra, L. Miao, D. G. Anderson, D. J. Crommelin, T. J. Anchordoquy, D. B. Volkin, W. Jiskoot, E. Mastrobattista, A. Wadhwa, A. Aljabbari, A. Lokras, C. Foged, A. Thakur, N. Fehrenbacher, and M. Jäättelä, "International Council for Harmonisation (ICH) Technical Requirements for Registration of Pharmaceuticals for Human Use, E6(R2) Good Clinical Practice (GCP) Guidance.," *Viruses*, vol. 12, no. 4, pp. 133–143, 2020.

[22] R. R. Goel, S. A. Apostolidis, M. M. Painter, D. Mathew, A. Pattekar, O. Kuthuru, S. Gouma, P. Hicks, W. Meng, A. M. Rosenfeld, S. Dysinger, K. A. Lundgreen, L. Kuri-Cervantes, S. Adamski, A. Hicks, S. Korte, D. A. Oldridge, A. E. Baxter, J. R. Giles, M. E. Weirick, C. M. McAllister, J. Dougherty, S. Long, K. D'Andrea, J. T. Hamilton, M. R. Betts, E. T. Luning Prak, P. Bates, S. E. Hensley, A. R. Greenplate, and E. J. Wherry, "Distinct antibody and memory B cell responses in SARS-CoV-2 naïve and recovered individuals following mRNA vaccination.," *Sci. Immunol.*, vol. 6, no. 58, pp. 1–20, 2021.

[23] M. V. Stankov, A. Cossmann, A. Bonifacius, A. Dopfer-Jablonka, G. M. Ramos, N. Gödecke, A. Z. Scharff, C. Happle, A.-L. Boeck, A. T. Tran, I. Pink, M. M. Hoeper, R. Blasczyk, M. S. Winkler, I. Nehlmeier, A. Kempf, H. Hofmann-Winkler, M. Hoffmann, B. Eiz-Vesper, S. Pöhlmann, and G. M. N. Behrens, "Humoral and Cellular Immune Responses Against Severe Acute Respiratory Syndrome Coronavirus 2 Variants and Human Coronaviruses After Single BNT162b2 Vaccination," *Clin. Infect. Dis.*, pp. 1–9, 2021.

[24] A. T. Widge and N. Rouphael, "Durability of Responses after SARS-CoV-2 mRNA-1273 Vaccination," *N. Engl. J. Med.*, vol. 384, no. 1, pp. 7–10, 2021.

[25] Z. Wang, F. Schmidt, Y. Weisblum, F. Muecksch, C. O. Barnes, S. Finkin, D. Schaefer-Babajew, M. Cipolla, C. Gaebler, J. A. Lieberman, T. Y. Oliveira, Z. Yang, M. E. Abernathy, K. E. Huey-Tubman, A. Hurley, M. Turroja, K. A. West, K. Gordon, K. G. Millard, V. Ramos, J. D. Silva, J. Xu, R. A. Colbert, R. Patel, J. Dizon, C. Unson-O'Brien, I. Shimeliovich, A. Gazumyan, M. Caskey, P. J. Bjorkman, R. Casellas, T. Hatziioannou,

P. D. Bieniasz, and M. C. Nussenzweig, “mRNA vaccine-elicited antibodies to SARS-CoV-2 and circulating variants,” *Nature*, vol. 592, no. April, 2021.

[26] M. S. Suthar, P. S. Arunachalam, M. Hu, N. Reis, and M. Trisal, “Durability of immune responses to the BNT162b2 mRNA vaccine,” 2021.

[27] J. Mateus, J. M. Dan, Z. Zhang, C. R. Moderbacher, M. Lammers, B. Goodwin, A. Sette, S. Crotty, and D. Weiskopf, “Low-dose mRNA-1273 COVID-19 vaccine generates durable memory enhanced by cross-reactive T cells,” *Science*, vol. 0, no. 0, p. eabj9853.
